# Lack of Evidence for a Reduced Late Positive Potential in Major Depressive Disorder

**DOI:** 10.1101/2020.04.29.20085571

**Authors:** Stevan Nikolin, Nicholas Chand, Donel Martin, Jacqueline Rushby, Colleen K. Loo, Tjeerd W. Boonstra

## Abstract

**Background:** Individuals with major depressive disorder (MDD) present with deficits in emotional reactivity. Conflicting models have been proposed to explain this effect. We sought to test the *emotional context insensitivity* hypothesis, which suggests that reactivity to positive and negatively-valenced emotional stimuli is blunted in depression, in a preregistered study.

**Methods:** Forty-one depressed participants and 41 age- and gender-matched healthy controls were presented a series of unpleasant and neutrally-valenced pictures in a passive view paradigm while acquiring electroencephalography (EEG). The late positive potential (LPP), an EEG correlate of emotional reactivity, was compared between groups using mixed-effects repeated-measures models and exploratory cluster-based permutation tests. A sensitivity analysis was performed to assess the robustness of LPP findings by reanalysing the LPPs using 22 EEG pipelines from studies identified in the literature.

**Results:** We found no difference in LPP amplitudes between MDD and healthy individuals using the preregistered analysis pipeline. The sensitivity analysis revealed that the magnitude and direction of LPP effect sizes were affected by the analysis pipeline. Exploratory permutation analyses revealed an electrode cluster that showed a significant reduction in the LPP for MDD participants while viewing unpleasant pictures.

**Conclusions:** These results do not provide evidence in support of the emotional context insensitivity hypothesis, except for the exploratory data-driven approach. Methodological differences, in particular in the analysis pipeline, contribute to the heterogeneity of LPP modulation in depression. A standardised approach to quantify EEG correlates of emotional reactivity is needed to evaluate alternative models of emotional reactivity in depression.

## INTRODUCTION

Major depressive disorder (MDD) is a mental illness that presents as persistent low mood and anhedonia, loss of interest in pleasurable activities, or both, in conjunction with other symptoms (American Psychiatric Association, 2013). Consistent with the core symptom of disturbed mood, depressed individuals report altered reactivity to emotion-eliciting contexts of positive or negative affect (Rottenberg et al., 2005). An improved understanding of the emotional deficits in MDD may thus provide further insight into mood symptoms (Rottenberg, 2005). Although the terms mood and emotion are sometimes used interchangeably, mood typically refers to slow-moving affective states that are weakly influenced by acute external stimuli, whereas emotions are considered to be rapid reactions to meaningful affective stimuli (Watson, 2000). Notwithstanding, the two are deeply intertwined and an improved understanding of the emotional deficits in MDD may thus provide further insight into mood symptoms (Rottenberg, 2005).

Three competing hypotheses have been proposed to describe the observed abnormalities in emotional reactivity in depressed individuals: 1) The *positive attenuation* model suggests that the physiological response is diminished to positive stimuli and unchanged for unpleasant, negatively-valenced stimuli (Clark and Watson, 1991), 2) The *negative potentiation* hypothesis states that depression increases reactivity towards negative contexts (Browning et al., 2010), and 3) The *emotional context insensitivity* model argues that depression results in blunted emotional reactivity to positive stimuli as well as to negative emotional contexts (Rottenberg et al., 2005). These theories disagree with regards to the physiological response in MDD to negative stimuli: *positive attenuation* predicts no difference in reactivity compared to healthy individuals, *negative potentiation* predicts increased reactivity, and the *emotional context insensitivity* predicts decreased reactivity in depression.

Emotional reactivity is commonly assessed using electroencephalography (EEG). Specifically, the late positive potential (LPP) may provide a reliable measure of affective cortical processing (Codispoti et al., 2006, 2007). The LPP is characterized as the mean amplitude of the event-related potential (ERP) within a time window approximately 400-1000ms following the visual presentation of emotionally laden images (Cuthbert et al., 2000; Schupp et al., 2000). Self-reported depressive symptoms have been associated with reduced LPP amplitudes to positive and unpleasant emotional stimuli in a non-clinical sample (Hill et al., 2019). Further, studies of depressed individuals have demonstrated attenuated LPP amplitudes in response to unpleasant stimuli (Benau et al., 2019; Foti et al., 2010; Grunewald et al., 2019; MacNamara et al., 2016), and pleasant pictures (Klawohn et al., 2020), in agreement with a summary of the LPP literature in a recent review by Hajcak and Foti (2020). However, these findings are not consistently observed, with some research reporting a significant increase in LPP amplitudes while passively observing unpleasant pictures (Auerbach et al., 2015; Burkhouse et al., 2017; Zhang et al., 2016), or no statistically significant difference (Weinberg et al., 2016). Although a meta-analysis found that MDD was characterized by reduced emotional reactivity to both positively and negatively valenced stimuli (Bylsma et al., 2008), in agreement with the *emotional context insensitivity* hypothesis, the results also demonstrated substantial heterogeneity between studies that could not be attributed to a specific moderator.

One of the potential contributing factors of heterogeneity between studies examining LPP amplitudes in response to emotional images is the role of emotion regulation in depression. In healthy individuals, the LPP is sensitive to the use of emotion regulation strategies, which refers to conscious or subconscious efforts to exert control over one’s emotional experience (Gross and John, 2003). Differences in the frequency or type of emotion regulation strategy use could partly explain the observed differences in the LPP to unpleasant images in MDD participants compared to healthy controls (Foti and Hajcak, 2008; Hajcak and Nieuwenhuis, 2006; Moser et al., 2009). In addition to emotion regulation strategy, anxiety, a common comorbidity of depression (Brown et al., 1998), has also been found to modulate the LPP. Individuals with high self-reported anxiety showed enhanced LPP amplitudes to aversive pictures compared to neutral pictures (MacNamara and Hajcak, 2009; MacNamara et al., 2016). Anxiety in depressed individuals may therefore contribute to the conflicting evidence for alternative models of emotional reactivity in MDD.

The primary aim of this study was therefore to verify previous reports in support of the *emotional context insensitivity* hypothesis by comparing the contrast in LPP amplitudes for unpleasant *vs* neutral pictures between MDD participants and healthy controls. We hypothesised that the LPP amplitude difference (i.e., unpleasant *vs* neutral) would be attenuated in MDD participants in support of *emotional context insensitivity*. We performed additional secondary analyses to examine whether emotion regulation strategy use and comorbid anxiety in MDD participants modulate the LPP amplitude in response to unpleasant pictures. We anticipated that MDD participants would use emotion regulation strategies less frequently than healthy controls, and that LPP amplitude would be moderated by emotion regulation strategy use and anxiety comorbidity in MDD participants. We based the research protocol used to test our hypotheses – the required sample size, use of affective picture stimuli, time interval, and region of interest to quantify the LPP amplitude – on previous literature, and documented this in a preregistration form (Nikolin et al., 2020). Preregistration provides a clear distinction between confirmatory and exploratory research and improves effect-size estimation, and may therefore help to evaluate the LPP as a biomarker for depression (Clayson et al., 2020). In addition, we identified 22 EEG processing pipelines reported in previous studies on the LPP in depression and reanalysed our data using these alternative processing pipelines to assess the robustness of the attenuation of the LPP amplitude difference to variations in processing parameters.

## MATERIALS AND METHODS

### Participants

A total of 82 participants were recruited in this study, of whom 41 were currently depressed and 41 were healthy age- and gender-matched controls. Individual participant data could only be obtained from Foti et al. (2010) to estimate the effect size for the contrast in LPP amplitudes between MDD participants and healthy controls for unpleasant *vs* neutral pictures. This showed a large effect size (Cohen’s *d* = 0.89), which would require 21 participants per diagnostic group to detect. We approximately doubled this sample size to allow for the detection of a more conservative, moderate effect size of Cohen’s *d* = 0.6 (using β = 80% and α = 0.05).

Inclusion criteria shared by both MDD participants and healthy controls were: aged 18 years and above, able to provide informed consent, normal or corrected-to-normal vision, no recent serious head injury in the last 12 months (e.g. loss of consciousness of more than 30 minutes), no current or past neurological condition (e.g. stroke, epilepsy), no history of drug or alcohol dependence in the last three months, not pregnant or possibly pregnant, no past history of distress reaction to affective images, and an ability to cooperate with the EEG procedure (e.g. no tremor).

Depressed participants were recruited from those seeking entry into clinical treatment trials at the Black Dog Institute, Sydney Australia. They were assessed by an experienced study Psychiatrist and confirmed to meet DSM-5 criteria for a current MDD diagnosis using the Structured Clinical Interview for DSM-5 Research Version (First et al., 2015), as well as score ≥ 20 on the Montgomery–Åsberg Depression Rating Scale for depressive symptoms (Montgomery and Asberg, 1979). Healthy controls were recruited via the University of New South Wales (UNSW) School of Psychology Paid Sign-Up System, Black Dog Institute Volunteer Research Register, or flyers distributed in the local area. Healthy control participants were eligible to participate if they had no current or past psychiatric or neurological disorders and were selected to be the same gender and within five years of age of the matched MDD participants since these factors have previously been shown to influence the latency, amplitude, and topography of ERP components (Kayser et al., 2000; Oliver-Rodriguez et al., 1999; Renfroe et al., 2016). The study protocol was approved by the UNSW Human Research Ethics Committee (HC16617).

### Procedure

Participants were instructed to passively view and attend to picture stimuli, selected from the International Affective Picture System (IAPS; Lang et al. (2008)). Pictures were chosen to be consistent with those used in previous EEG studies investigating LPP effects (Hajcak and Nieuwenhuis, 2006; MacNamara et al., 2016; Moser et al., 2006; Weinberg et al., 2016). Participants viewed 56 images once each, of which 28 were unpleasant (e.g., threatening scenes, violence, weapons) and 28 were neutral (e.g., neutral faces, household objects, landscapes). The normative valence ratings were lower for unpleasant pictures (2.79 ± 0.59) than for neutral pictures (5.02 ± 0.20), indicating that they elicited more negative emotional affect. Normative arousal ratings were higher for unpleasant pictures (5.74 ± 0.84) than for neutral pictures (2.98 ± 0.59; see Supplementary Materials for further details).

The stimuli were presented on a 58.4 cm (23-inch) computer screen in a dimly lit, sound-attenuated room using Inquisit 4 software (Version 4, Millisecond Software). Digitised pictures sized 20 × 30 cm were presented on a black background, spanning a visual angle of 20-30 degrees, consistent with previous EEG studies (Dunning and Hajcak, 2009; MacNamara et al., 2016). The pictures were displayed for 5 s in a pre-determined randomised order, so participants were unable to anticipate the valence of each picture. A black screen with a centered white fixation cross was presented for 2 s between stimuli (Fig. 1). Pictures of the same valence were not presented more than three times consecutively to prevent participants from generating expectations of upcoming stimuli and to avoid over-saturation of emotional salience. Participants were not informed of any emotional regulation strategies prior to viewing the IAPS images.

**Figure 1.**
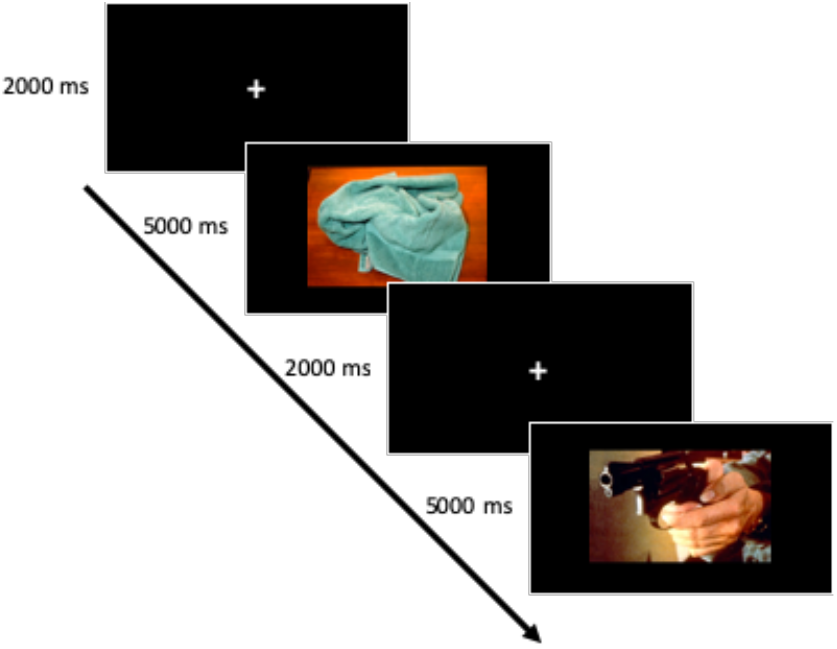
Study protocol. Participants passively observed a total of 56 picture stimuli. These were presented in a randomised order and consisted of 28 unpleasant images with a negative valence, and 28 neutral images with no emotional valence, sourced from the International Affective Picture System (IAPS). Pictures were displayed for 5 s, followed by a 2 s fixation cross. A sample of two trials is displayed, showing a neutral image (a towel) and an unpleasant image (a gun).

### Behavioural measures

Prior to the experimental session, MDD participants had their mood assessed by a trained rater using the ten-item Montgomery-Åsberg Depression Rating Scale (MADRS) for depressive symptoms (Montgomery and Asberg, 1979). All participants completed the 21-item self-reported Depression, Anxiety and Stress Scale (DASS; Lovibond and Lovibond (1995)). The DASS was chosen as it reliably discriminates between depression, anxiety, and stress states (Brown et al., 1997; Crawford and Henry, 2003; Henry and Crawford, 2005). Upon completion of the experimental session, we used an emotion regulation questionnaire adapted from Gross and John (2003), provided in Supplementary Materials. Participants were able to select from five emotion regulation strategies, including no strategy, reinterpretation, distraction, detachment, and ‘other’ in which participants could specify their strategy. Strategies specified in the ‘other’ category were re-assigned to one of the former strategy options by two study personnel who were blind to participant group allocation.

### Electroencephalography data acquisition and processing

EEG data were acquired using a 64-channel setup and a TMSi Refa amplifier (TMS International, Oldenzaal, Netherlands) with a sampling rate of 1024 Hz. Offline EEG processing and analyses were conducted using custom-developed MATLAB scripts (v.R2019a; MathWorks) and the Fieldtrip toolbox (Oostenveld et al., 2011). All scripts used for EEG processing and calculation of neurophysiological measures are available at the following link: https://github.com/snikolin/MDDvsCTRL_IAPS.

EEG data were pre-processed using a Butterworth IIR digital filter to remove 50 Hz electrical line noise. Following this, a second-order band-pass filter with cut-offs of 0.5 and 70 Hz was used to remove low and high frequency noise generated from head movements and muscle activity. The EEG data were then epoched into 6 s intervals, beginning 1 s before each picture onset and continuing for 5 s after picture onset. Epochs were rejected firstly using a semi-automated procedure, and then on visual inspection to identify any remaining trials with large artefacts. Smaller, non-cortical physiological activity (e.g., cardiac, muscle, or ocular) and non-physiological activity (e.g., environmental noise, or movement) were removed using an Independent Components Analysis (ICA) algorithm (Delorme et al., 2007; Hyvärinen et al., 2001; Makeig et al., 1996). Finally, EEG data was re-referenced to the common average reference, which reduces the distortion of measured EEG potentials as compared to a linked mastoids reference (Hu et al., 2018).

#### Event-related potentials

ERPs were constructed by separately averaging trials for neutral and unpleasant pictures time-locked to picture presentation. Each averaged ERP was baseline corrected to the mean activity occurring in the 200 ms time interval prior to stimulus onset. The late positive potential (LPP) is a slow, positive wave occurring maximally at centroparietal recording sites, beginning approximately 400 ms after stimulus onset and lasting several seconds (Cuthbert et al., 2000; Schupp et al., 2000). We quantified the LPP component as the mean amplitude of the ERP from 400 to 1000 ms following stimulus onset at a centroparietal cluster of EEG channels (Cz, CPz, Pz, CP1, and CP2), where it has previously been shown to be maximal (Foti et al., 2010; MacNamara et al., 2016). Participants with LPP component amplitudes greater than three standard deviations from the mean were considered outliers and excluded from statistical analyses.

### Statistical analyses

For the primary analysis, LPPs were investigated using mixed-effects repeated-measures model (MRMM), performed using SPSS software (IBM SPSS Statistics 25, SPSS Inc.), and were in accordance with our preregistered protocol (Nikolin et al., 2020). The model included a repeated factor of Valence (unpleasant vs neutral) using an unstructured covariance matrix, as well as the between-subjects factor of Group (MDD vs healthy controls), and the Valence × Group interaction. Subject was included as a random effect.

Residuals were visually inspected to ensure an adequate fit of the model. Cohen’s *d* was calculated to estimate the effect size for the LPP amplitude interaction contrast between unpleasant and neutral images comparing MDD participants to healthy controls:

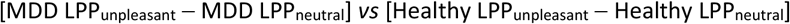

A follow-up exploratory non-parametric cluster-based permutation test was performed using the Fieldtrip toolbox in MATLAB to identify any significant differences in emotional stimulus processing between MDD participants and healthy controls beyond the *a priori* time interval and channels operationalised for the LPP. This method controls for multiple comparisons while comparing differences across a large spatiotemporal parameter space (Maris and Oostenveld, 2007). Participant ERPs, incorporating all EEG channels within the time interval 0 – 1000 ms following presentation of IAPS stimuli, were randomly assigned to one of two group conditions in the same proportion as the original data for 3000 permutations. The permuted data were compared using independent samples *t*-tests, producing a data-driven distribution of the test statistics. The *t*-test statistic for the original, non-permuted data was then compared against the permuted distributions to identify any significant clusters. A value of α < 0.05 was adopted as the two-tailed significance threshold. Statistically significant clusters were required to comprise at least two neighbouring channels.

For secondary analyses, a Fisher’s Exact Test was used to investigate whether there were differences in emotion regulation strategy use between diagnostic groups. Emotion regulation strategies were entered into a 2 × 2 contingency table using the binary variables of Strategy (no strategy vs strategy) and Group (MDD vs healthy controls). Emotional regulation strategies consisted of reinterpretation, distraction, or detachment.

The effects of depression on LPP amplitude may become more apparent when accounting for variability in emotion regulation strategy use or comorbid anxiety symptoms across diagnostic groups. To this end, we repeated MRMM analyses with emotion regulation strategy or comorbid anxiety entered separately as covariates. Strategy was represented as a binary categorical variable (no strategy *vs* strategy). Anxiety was included as a continuous variable using the anxiety subscore of the DASS. Additional confirmatory simple univariate linear regression analyses were performed with LPP amplitude as the dependent variable, restricted to amplitudes obtained from MDD participants while viewing unpleasant images.

Scripts for EEG analyses in MATLAB and statistical analyses using SPSS are provided at: https://github.com/snikolin/MDDvsCTRL_IAPS.

### Sensitivity analysis

Given the variability in the methodology used in previous studies to quantify the LPP, we performed additional analyses to assess the robustness of our findings and recomputed the LPP using pipelines identified in the depression literature. We performed a PubMed review on the 8^th^ of May 2021 for manuscripts containing the terms: depression AND (electroencephalography OR EEG) AND “late positive potential”. Studies were included if they reported LPP results in participants with depression or depressive tendencies and provided a comprehensive description of their EEG processing pipeline. We supplemented the review with additional studies identified while reading this literature. We also assessed alternative variants of our preregistered pipeline that were not observed in the literature but may be of interest. These included: Repeating our preregistered pipeline analysis using linked mastoid re-referencing (Alternative #1) and reducing the high-pass filter to 0.1Hz (Alternative #2). We used a collapsed localiser approach (Luck and Gaspelin, 2017), which entails calculating the grand-average ERP for all participants (MDD and controls combined) for unpleasant relative to neutral IAPS images and using the resultant topography to select a sample-specific region of interest for the LPP (Alternative #3). Lastly, we repeated Alternative #2 using an average common reference (Alternative #4), and similarly repeated Alternative #3 using an average common reference (Alternative #5). We estimated effect sizes for each pipeline using Cohen’s *d*, as described previously. We additionally report significance *p*-values for simple independent samples *t*-tests for each pipeline’s contrast comparison (i.e., MDD LPP_unpleasant_ -MDD LPP_neutral_ *vs* Healthy LPP_unpleasant_ -Healthy LPP_neutral_). In total, we assessed 22 unique EEG processing pipelines and MRMMs were repeated using LPP amplitudes obtained from each pipeline.

Lastly, we repeated exploratory non-parametric cluster-based permutation tests using 1) a 0.5Hz high-pass filter with referencing to linked mastoids; 2) a 0.1Hz high-pass filter and common average reference; and lastly 3) a 0.1Hz high-pass filter with referencing to linked mastoids.

## RESULTS

Behavioural and EEG measures were assessed in 82 participants (41 with MDD and 41 aged- and gender-matched controls). Table 1 shows baseline demographic and clinical information for MDD participants and healthy controls. One participant was excluded from EEG analyses due to values greater than three standard deviations from the group mean, leaving a total of 40 MDD participants and 41 healthy controls. Following rejection of epochs with EEG artefacts, MDD participants had an average of 25.5 unpleasant trials (SD=2.1; range=20-28) and 26.1 neutral trials (SD=1.7; range=21-28). Healthy controls had an average of 25.8 unpleasant trials (SD=1.8; range=20-28) and 26.4 neutral trials (SD=1.5; range=21-28).

**Table 1.**
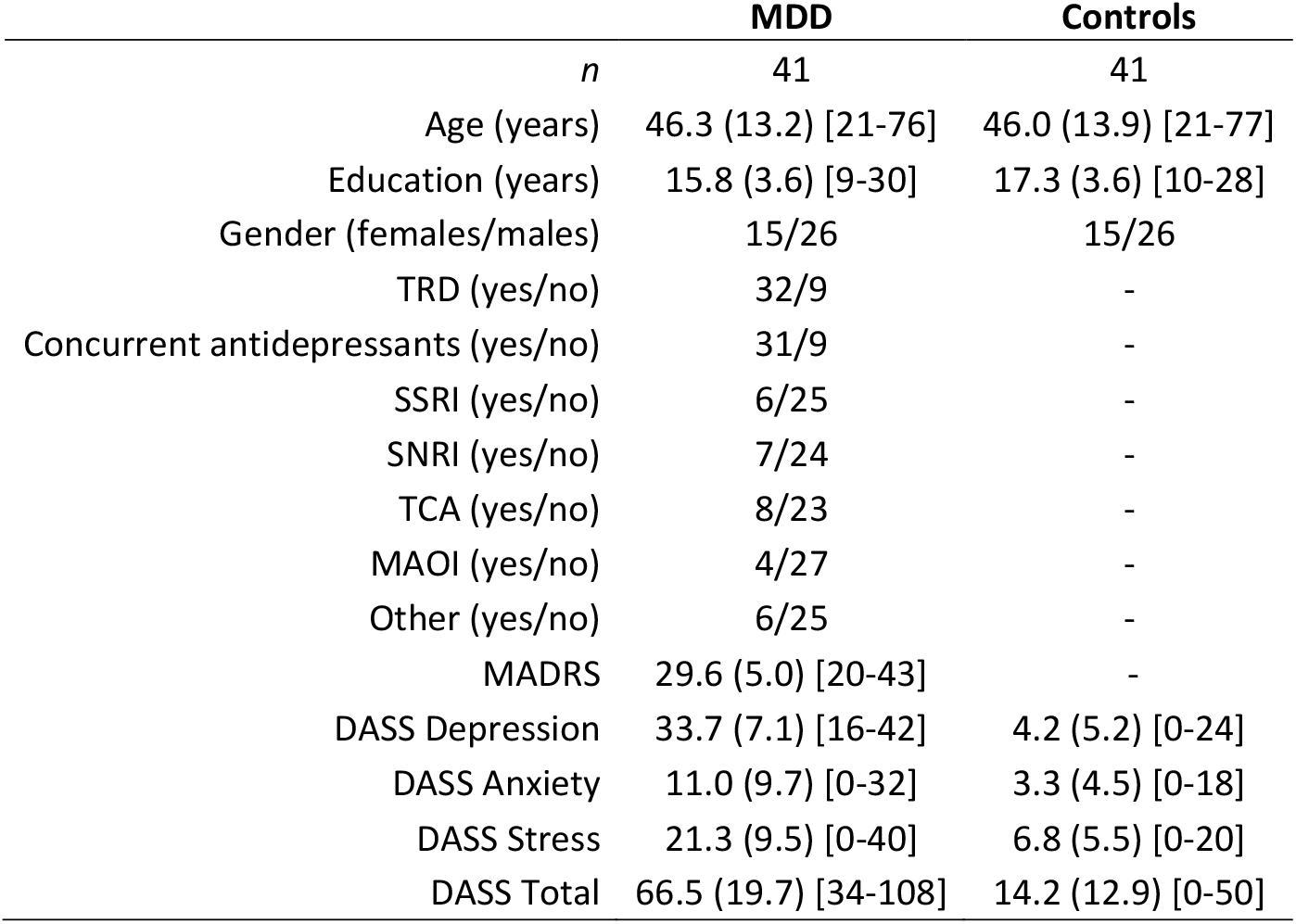
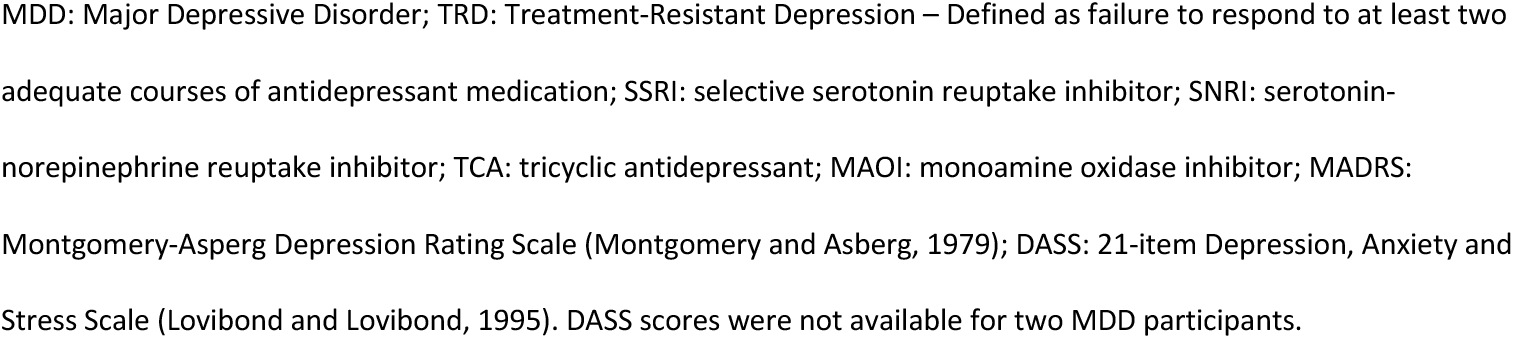
Baseline demographic and clinical information. Means, standard deviations (SD), and ranges [minimum-maximum] are shown for continuous measures, and frequency counts for categorical measures.

|  | **MDD** | **Controls** |
| --- | --- | --- |
| *n* | 41 | 41 |
| Age (years) | 46.3 (13.2) [21-76] | 46.0 (13.9) [21-77] |
| Education (years) | 15.8 (3.6) [9-30] | 17.3 (3.6) [10-28] |
| Gender (females/males) | 15/26 | 15/26 |
| TRD (yes/no) | 32/9 | - |
| Concurrent antidepressants (yes/no) | 31/9 | - |
| SSRI (yes/no) | 6/25 | - |
| SNRI (yes/no) | 7/24 | - |
| TCA (yes/no) | 8/23 | - |
| MAOI (yes/no) | 4/27 | - |
| Other (yes/no) | 6/25 | - |
| MADRS | 29.6 (5.0) [20-43] | - |
| DASS Depression | 33.7 (7.1) [16-42] | 4.2 (5.2) [0-24] |
| DASS Anxiety | 11.0 (9.7) [0-32] | 3.3 (4.5) [0-18] |
| DASS Stress | 21.3 (9.5) [0-40] | 6.8 (5.5) [0-20] |
| DASS Total | 66.5 (19.7) [34-108] | 14.2 (12.9) [0-50] |

MDD: Major Depressive Disorder; TRD: Treatment-Resistant Depression – Defined as failure to respond to at least two adequate courses of antidepressant medication; SSRI: selective serotonin reuptake inhibitor; SNRI: serotonin-norepinephrine reuptake inhibitor; TCA: tricyclic antidepressant; MAOI: monoamine oxidase inhibitor; MADRS: Montgomery-Asberg Depression Rating Scale (Montgomery and Asberg, 1979); DASS: 21-item Depression, Anxiety and Stress Scale (Lovibond and Lovibond, 1995). DASS scores were not available for two MDD participants.

### Primary Analysis

The LPP amplitude was compared across groups (MDD and controls) and valence conditions (unpleasant and neutral). MRMM results showed a significant main effect of Valence (F = 19.62, *p* < 0.001), indicating increased LPP amplitude for unpleasant compared to neutral images (Fig. 2). The main effect of Group (F = 0.39, *p* = 0.54) and the Group × Valence interaction (F = 0.26, *p* = 0.61) were not significant. Comparison of the valence difference between MDD participants and healthy controls showed a small effect size (*d* = -0.11).

**Figure 2.**
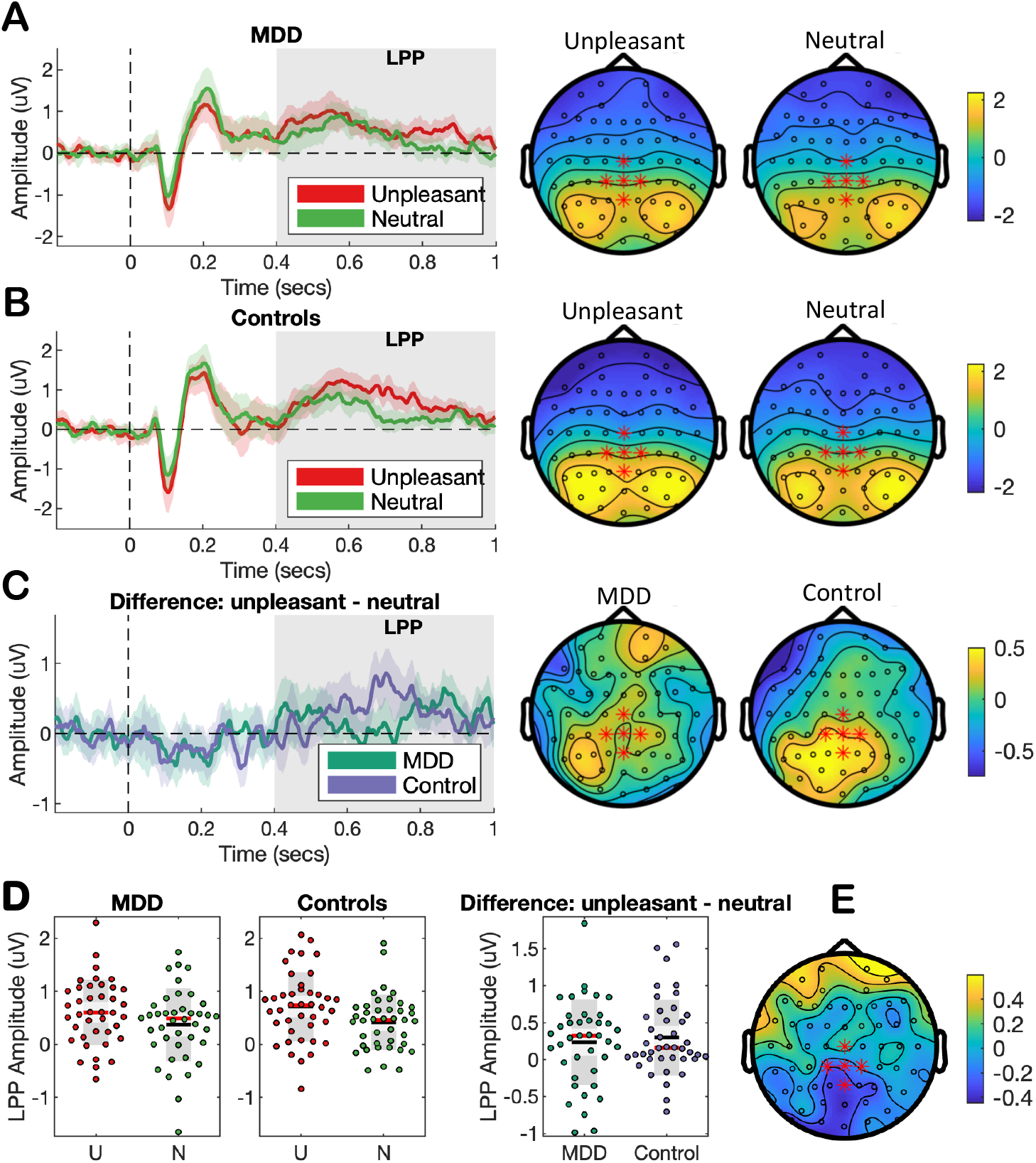
Preregistered EEG measures of affective processing. The LPP was quantified as the mean EEG amplitude in the time interval from 400 to 1000 ms following stimulus onset for a cluster of centroparietal EEG channels (highlighted in red). ERPs, spatial topographies, and scatter plots of LPP amplitude are presented for MDD participants and controls while viewing unpleasant and neutral images. ERPs are displayed with bootstrapped 95% confidence intervals and with the time interval used to calculate LPPs shaded in light grey. Spatial topographies are of average ERP amplitude from 400 to 1000 ms. Scatter plots show the mean (black line), median (red line), and the 95% confidence interval (dark grey region). **A)** ERPs and topographies for MDD participants. **B)** ERPs and topographies for healthy controls. **C)** ERPs and spatial topographies of the valence difference (unpleasant – neutral) for MDD participants and healthy controls. **D)** Scatter plots of LPP amplitude for unpleasant (U) and neutral (N) images at the centroparietal cluster. **E)** Spatial topography of the valence difference (unpleasant – neutral) contrasting MDD participants with healthy controls.

Exploratory non-parametric cluster-based permutation testing compared the difference scores (unpleasant – neutral stimuli) between MDD participants and healthy controls. These revealed a cluster showing a significant difference between MDD participants and controls (*p* = 0.01; Fig. 3). The time interval for the cluster fell within the preregistered LPP window (662 – 755 ms). EEG channels identified in this cluster were slightly more posterior than those for our preregistered LPP analysis and included CPz, CP1, Pz, POz, P1, and P2. Three of these channels are also found in the preregistered cluster (CPz, CP1, and Pz). See Supplementary Material for further details regarding cluster channels.

**Figure 3.**
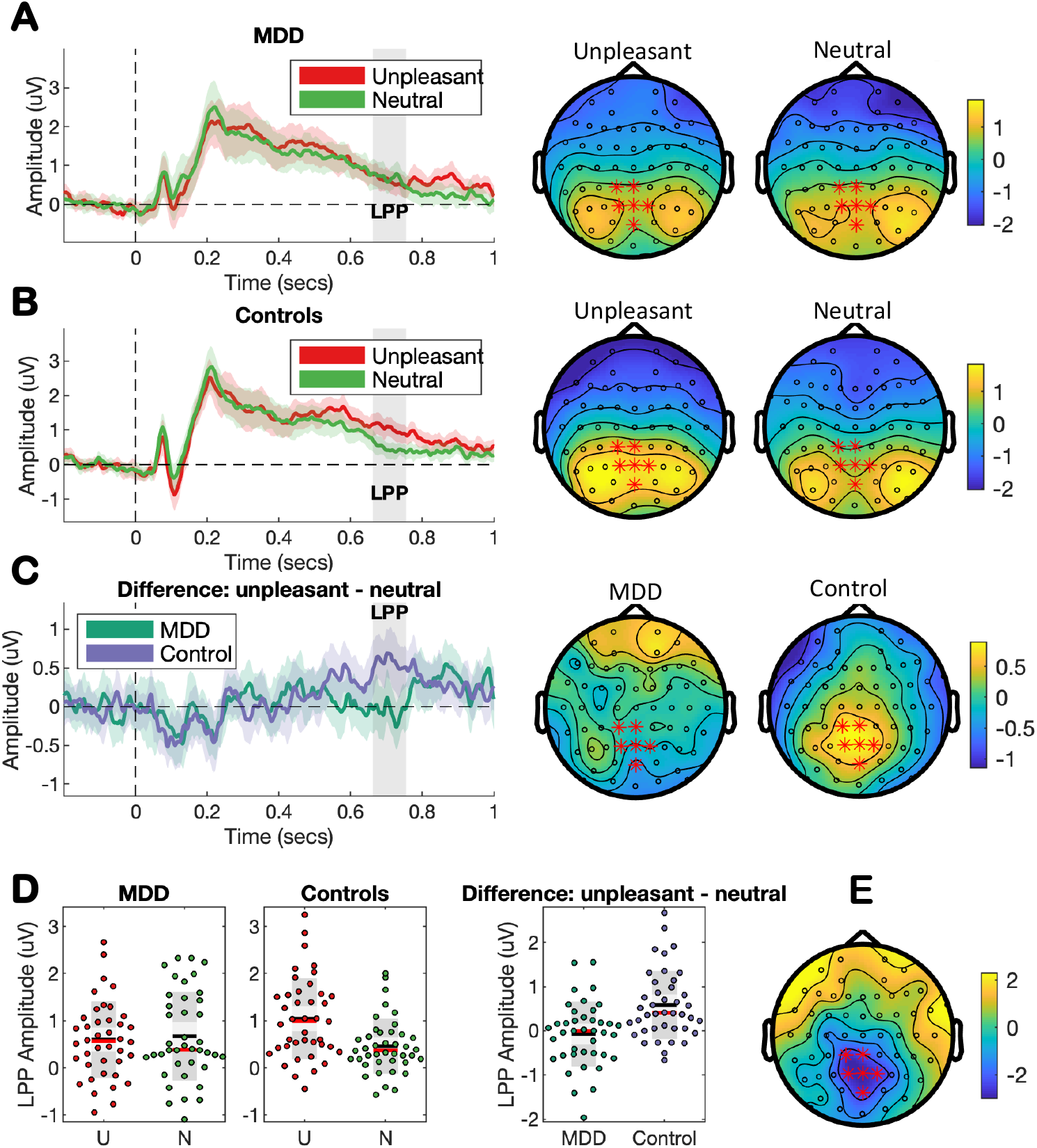
Exploratory EEG measures of affective processing. Exploratory cluster-based permutation testing revealed a significant cluster from 662 to 755 ms in EEG channels CPz, CP1, Pz, POz, P1, and P2 (highlighted in red). ERPs, spatial topographies, and scatter plots of LPP amplitude are presented for MDD participants and controls while viewing unpleasant and neutral images. ERPs are displayed with bootstrapped 95% confidence intervals and with the time interval used to calculate the mean amplitude within the exploratory cluster shaded in light grey. Spatial topographies are of average ERP amplitude from 662 to 755 ms. Scatter plots show the mean (black line), median (red line), and the 95% confidence interval (dark grey region). **A)** ERPs and topographies for MDD participants. **B)** ERPs and topographies for healthy controls. **C)** ERPs and spatial topographies of the valence difference (unpleasant – neutral) for MDD participants and healthy controls. **D)** Scatter plots of LPP amplitude for unpleasant (U) and neutral (N) images at the exploratory cluster. Comparison of the valence differences between MDD participants and healthy controls showed a large effect size of Cohen’s *d* = -0.88. **E)** Spatial topography of the valence difference (unpleasant – neutral) for the time interval 662 – 755 ms contrasting MDD participants with healthy controls.

### Secondary Analyses

As part of our secondary confirmatory analyses, we also investigated the association between strategy use and anxiety on the LPP amplitude. We compared the emotion regulation strategies used by MDD participants and control. Most participants reported that they did not use a strategy (Table 2). A Fisher’s Exact Test revealed that strategy use was not significantly different between both groups (p = 0.34).

**Table 2.** Emotion regulation strategy use.

|  | **MDD**<br>**(*n* = 40)** | **Controls**<br>**(*n* = 41)** |
| --- | --- | --- |
| *No strategy* | 30 | 26 |
| *Strategy* | 10 | 15 |
| Reinterpretation | 1 | 5 |
| Distraction | 2 | 5 |
| Detachment | 7 | 5 |

MDD: Major Depressive Disorder

We performed MRMMs with the inclusion of either Anxiety or Strategy as covariates. Neither the model including Strategy (F = 0.2, *p* = 0.67), nor the model including Anxiety (F = 0.2, *p* =0.63) were substantively changed (see Supplementary Materials). Next, we performed simple univariate linear regression analyses, restricting the dataset to LPP values obtained from MDD participants while viewing unpleasant images. Strategy was not significantly associated with LPP amplitudes (r = 0.13, r^2^ = 0.02, β = -0.18, p = 0.44; Fig. S1A). Similarly, Anxiety was not significantly associated with the LPP (r = 0.21, r^2^ = 0.05, β = 0.01, p = 0.20; Fig. S1B).

### Sensitivity analysis

We reanalysed our data using 22 alternative EEG processing pipelines (Table 3). A literature review identified 18 studies and the most used parameters were a high-pass filter of 0.1Hz (k=9), low-pass filter of 30Hz (k=16), correction for ocular artefacts using a regression method such as that proposed by Gratton et al., (1983) (k=14), and re-referencing to linked mastoids (k=12). The LPP time of interest was most often operationalised as starting at either 400 or 600 ms (k=7), and ending at 1000 ms (k=9). The region of interest was typically centro-parietal, mostly consisting of channels Pz (k=11), Cz (k=9), CP1 (k=9), and CP2 (k=9).

**Table 3.**
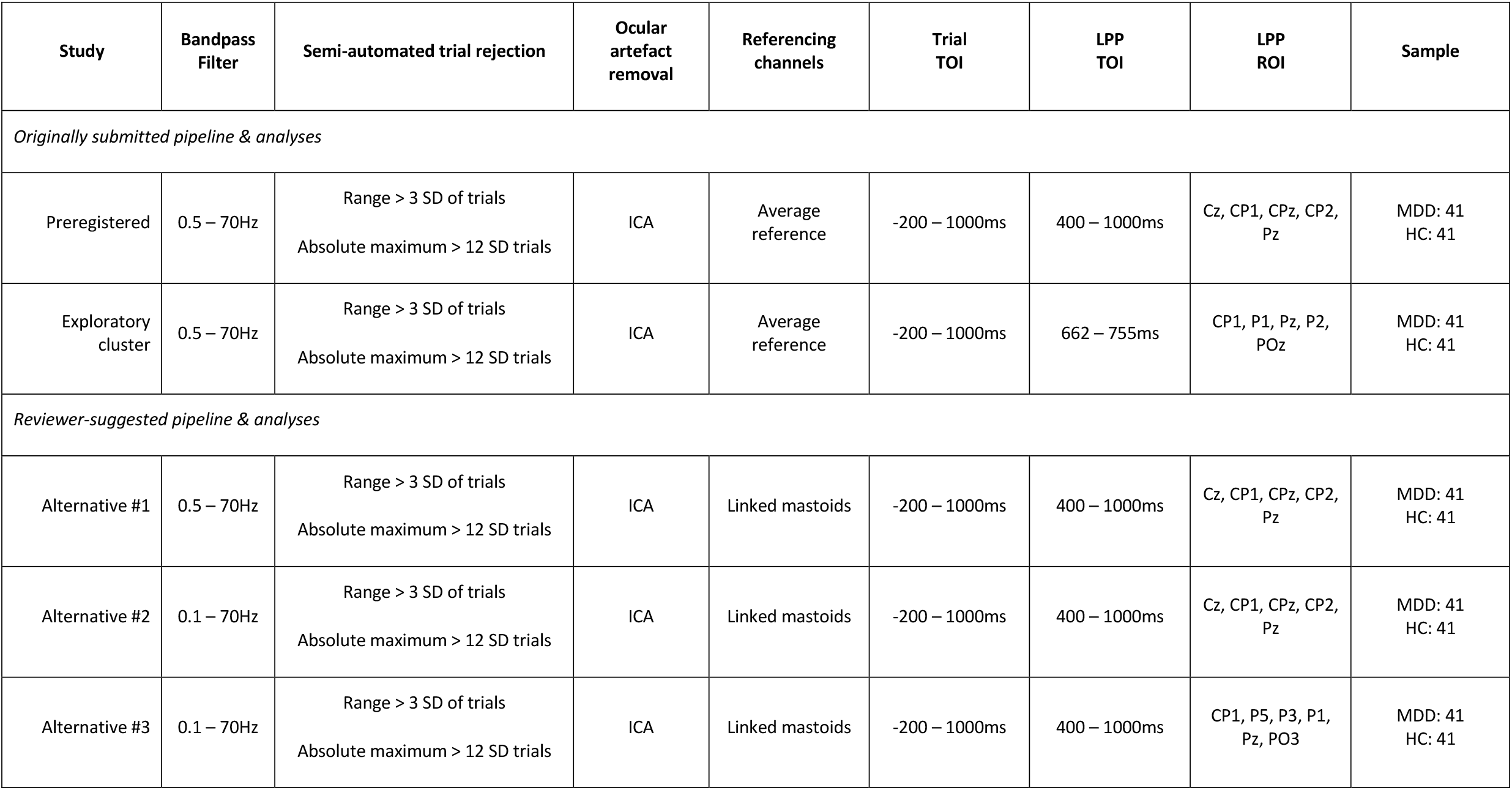
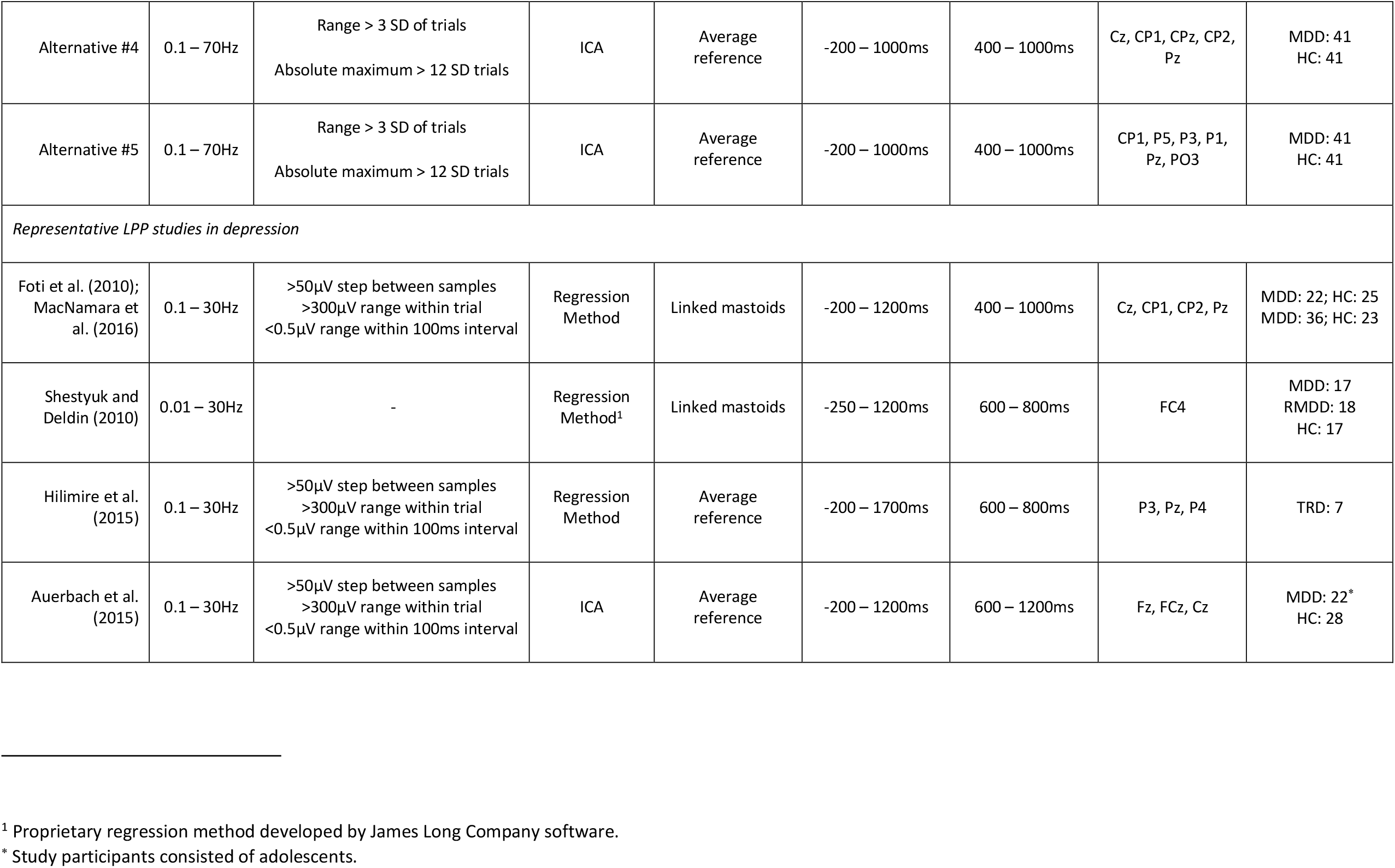
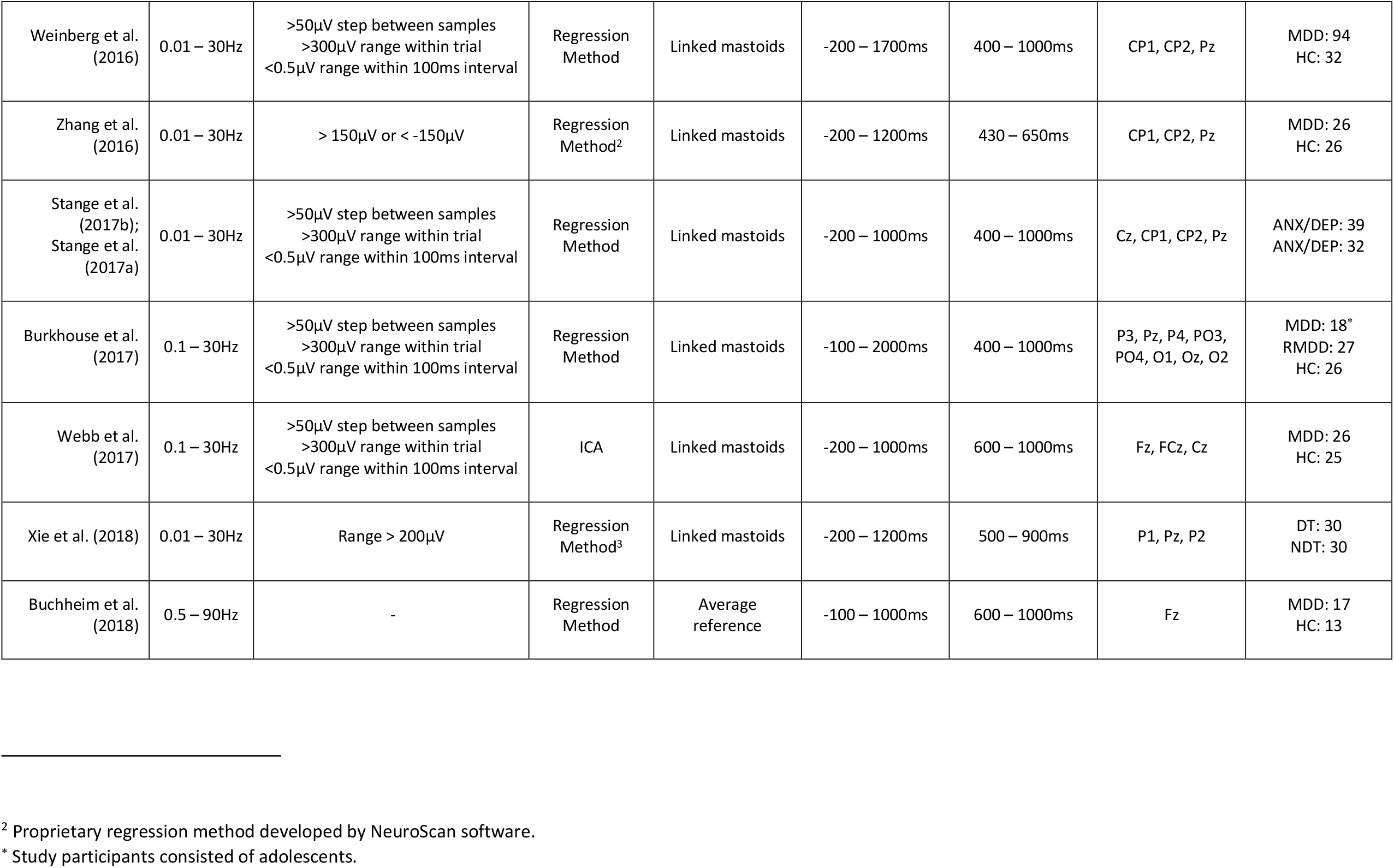
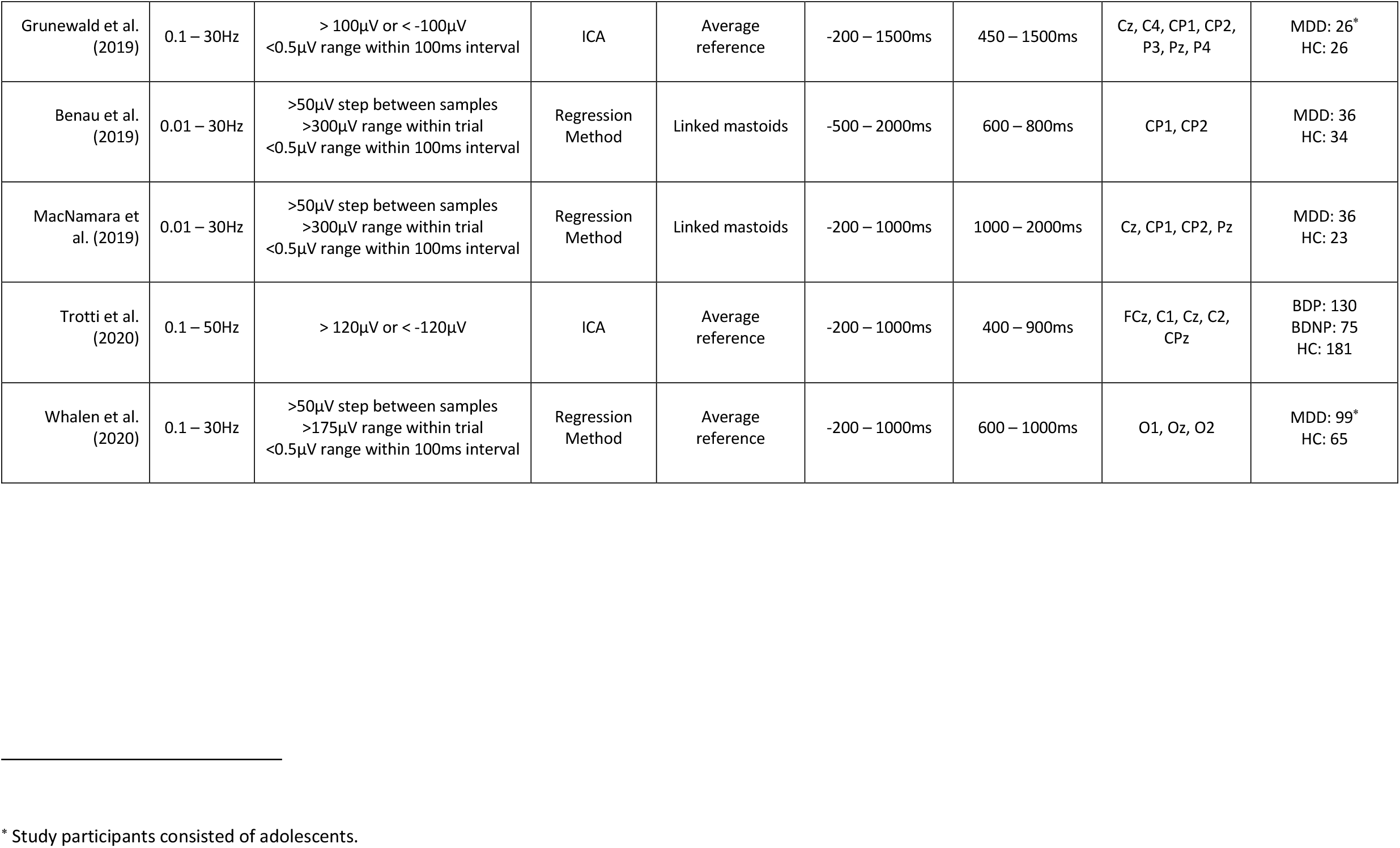
EEG pipeline processing steps.

| Study | Bandpass Filter | Semi-automated trial rejection | Ocular artefact removal | Referencing channels | Trial TOI | LPP TOI | LPP ROI | Sample |
| --- | --- | --- | --- | --- | --- | --- | --- | --- |
| *Originally submitted pipeline & analyses* |  |  |  |  |  |  |  |  |
| Preregistered | 0.5 – 70Hz | Range > 3 SD of trials<br>Absolute maximum > 12 SD trials | ICA | Average reference | -200 – 1000ms | 400 – 1000ms | Cz, CP1, CPz, CP2, Pz | MDD: 41<br>HC: 41 |
| Exploratory cluster | 0.5 – 70Hz | Range > 3 SD of trials<br>Absolute maximum > 12 SD trials | ICA | Average reference | -200 – 1000ms | 662 – 755ms | CP1, P1, Pz, P2, POz | MDD: 41<br>HC: 41 |
| *Reviewer-suggested pipeline & analyses* |  |  |  |  |  |  |  |  |
| Alternative #1 | 0.5 – 70Hz | Range > 3 SD of trials<br>Absolute maximum > 12 SD trials | ICA | Linked mastoids | -200 – 1000ms | 400 – 1000ms | Cz, CP1, CPz, CP2, Pz | MDD: 41<br>HC: 41 |
| Alternative #2 | 0.1 – 70Hz | Range > 3 SD of trials<br>Absolute maximum > 12 SD trials | ICA | Linked mastoids | -200 – 1000ms | 400 – 1000ms | Cz, CP1, CPz, CP2, Pz | MDD: 41<br>HC: 41 |
| Alternative #3 | 0.1 – 70Hz | Range > 3 SD of trials<br>Absolute maximum > 12 SD trials | ICA | Linked mastoids | -200 – 1000ms | 400 – 1000ms | CP1, P5, P3, P1, Pz, PO3 | MDD: 41<br>HC: 41 |
| Alternative #4 | 0.1 – 70Hz | Range > 3 SD of trials<br>Absolute maximum > 12 SD trials | ICA | Average reference | -200 – 1000ms | 400 – 1000ms | Cz, CP1, CPz, CP2, Pz | MDD: 41<br>HC: 41 |
| Alternative #5 | 0.1 – 70Hz | Range > 3 SD of trials<br>Absolute maximum > 12 SD trials | ICA | Average reference | -200 – 1000ms | 400 – 1000ms | CP1, P5, P3, P1, Pz, PO3 | MDD: 41<br>HC: 41 |
| *Representative LPP studies in depression* |  |  |  |  |  |  |  |  |
| Foti et al. (2010);<br>MacNamara et al. (2016) | 0.1 – 30Hz | >50µV step between samples<br>>300µV range within trial<br><0.5µV range within 100ms interval | Regression Method | Linked mastoids | -200 – 1200ms | 400 – 1000ms | Cz, CP1, CP2, Pz | MDD: 22; HC: 25<br>MDD: 36; HC: 23 |
| Shestyuk and Deldin (2010) | 0.01 – 30Hz | - | Regression Method<sup>1</sup> | Linked mastoids | -250 – 1200ms | 600 – 800ms | FC4 | MDD: 17<br>RMDD: 18<br>HC: 17 |
| Hilimire et al. (2015) | 0.1 – 30Hz | >50µV step between samples<br>>300µV range within trial<br><0.5µV range within 100ms interval | Regression Method | Average reference | -200 – 1700ms | 600 – 800ms | P3, Pz, P4 | TRD: 7 |
| Auerbach et al. (2015) | 0.1 – 30Hz | >50µV step between samples<br>>300µV range within trial<br><0.5µV range within 100ms interval | ICA | Average reference | -200 – 1200ms | 600 – 1200ms | Fz, FCz, Cz | MDD: 22*<br>HC: 28 |
| Weinberg et al. (2016) | 0.01 – 30Hz | >50μV step between samples<br>>300μV range within trial<br><0.5μV range within 100ms interval | Regression Method | Linked mastoids | -200 – 1700ms | 400 – 1000ms | CP1, CP2, Pz | MDD: 94<br>HC: 32 |
| Zhang et al. (2016) | 0.01 – 30Hz | > 150μV or < -150μV | Regression Method<sup>2</sup> | Linked mastoids | -200 – 1200ms | 430 – 650ms | CP1, CP2, Pz | MDD: 26<br>HC: 26 |
| Stange et al. (2017b);<br>Stange et al. (2017a) | 0.01 – 30Hz | >50μV step between samples<br>>300μV range within trial<br><0.5μV range within 100ms interval | Regression Method | Linked mastoids | -200 – 1000ms | 400 – 1000ms | Cz, CP1, CP2, Pz | ANX/DEP: 39<br>ANX/DEP: 32 |
| Burkhouse et al. (2017) | 0.1 – 30Hz | >50μV step between samples<br>>300μV range within trial<br><0.5μV range within 100ms interval | Regression Method | Linked mastoids | -100 – 2000ms | 400 – 1000ms | P3, Pz, P4, PO3,<br>PO4, O1, Oz, O2 | MDD: 18*<br>RMDD: 27<br>HC: 26 |
| Webb et al. (2017) | 0.1 – 30Hz | >50μV step between samples<br>>300μV range within trial<br><0.5μV range within 100ms interval | ICA | Linked mastoids | -200 – 1000ms | 600 – 1000ms | Fz, FCz, Cz | MDD: 26<br>HC: 25 |
| Xie et al. (2018) | 0.01 – 30Hz | Range > 200μV | Regression Method<sup>3</sup> | Linked mastoids | -200 – 1200ms | 500 – 900ms | P1, Pz, P2 | DT: 30<br>NDT: 30 |
| Buchheim et al. (2018) | 0.5 – 90Hz | - | Regression Method | Average reference | -100 – 1000ms | 600 – 1000ms | Fz | MDD: 17<br>HC: 13 |
| Grunewald et al. (2019) | 0.1 – 30Hz | > 100µV or < -100µV<br><0.5µV range within 100ms interval | ICA | Average reference | -200 – 1500ms | 450 – 1500ms | Cz, C4, CP1, CP2, P3, Pz, P4 | MDD: 26*<br>HC: 26 |
| Benau et al. (2019) | 0.01 – 30Hz | >50µV step between samples<br>>300µV range within trial<br><0.5µV range within 100ms interval | Regression Method | Linked mastoids | -500 – 2000ms | 600 – 800ms | CP1, CP2 | MDD: 36<br>HC: 34 |
| MacNamara et al. (2019) | 0.01 – 30Hz | >50µV step between samples<br>>300µV range within trial<br><0.5µV range within 100ms interval | Regression Method | Linked mastoids | -200 – 1000ms | 1000 – 2000ms | Cz, CP1, CP2, Pz | MDD: 36<br>HC: 23 |
| Trotti et al. (2020) | 0.1 – 50Hz | > 120µV or < -120µV | ICA | Average reference | -200 – 1000ms | 400 – 900ms | FCz, C1, Cz, C2, CPz | BDP: 130<br>BDNP: 75<br>HC: 181 |
| Whalen et al. (2020) | 0.1 – 30Hz | >50µV step between samples<br>>175µV range within trial<br><0.5µV range within 100ms interval | Regression Method | Average reference | -200 – 1000ms | 600 – 1000ms | O1, Oz, O2 | MDD: 99*<br>HC: 65 |

\* Study participants consisted of adolescents.

We repeated our primary MRMM analyses using the pipelines described in Table 3 with the results provided in Supplementary Table S2. In all pipelines where a regression was used to remove the ocular artefact we implemented the procedure as outlined in Gratton et al. (1983), as this was used most often and contained sufficient information for replication. Statistical findings remained qualitatively similar for these alternative analysis approaches. Briefly, all pipelines showed a significant effect of Valence, with the exception of Whalen et al. (2020). Two pipelines resulted in a significant effect of Group: in one pipeline the MDD LPPs were more negative than controls (Hilimire et al., 2015), the other pipeline resulted in the opposite effect of MDD LPPs being more positive than controls (Buchheim et al., 2018), although in both cases depressed LPP amplitude was attenuated. No processing pipeline resulted in a significant Group × Valence interaction effect (all *p* > 0.05). Bootstrapped Cohen’s *d* effect sizes ranged from 0.3 (in the direction of *negative potentiation* in depression) to -0.4 (in the direction of *emotional context insensitivity*); however, these were not statistically significant (Fig. 4).

**Figure 4.**
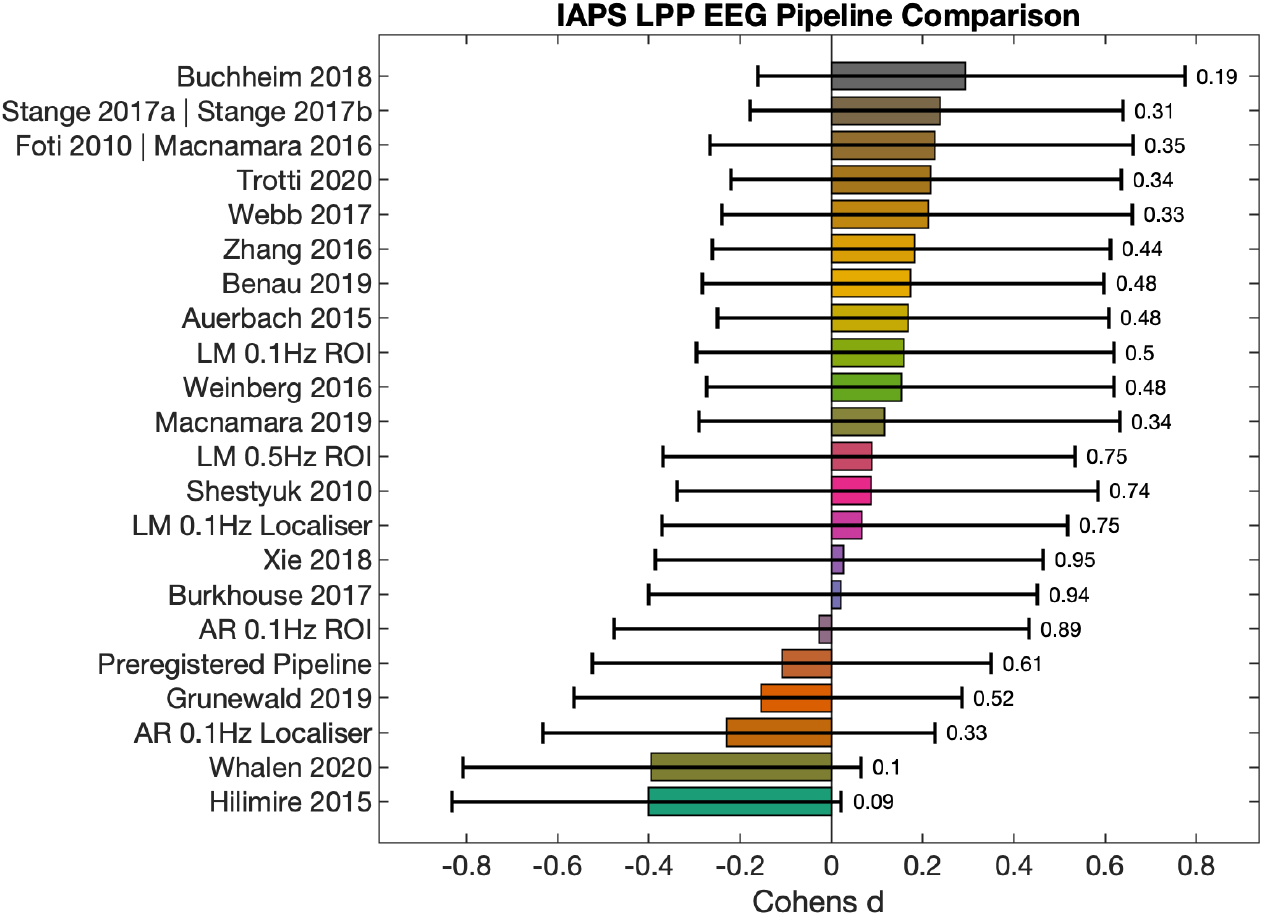
Effect sizes of 22 alternative EEG processing pipelines. Bootstrapped Cohen’s *d* effect sizes with 95% confidence intervals were calculated for the interaction contrast between unpleasant and neutral images comparing MDD participants to healthy controls: [MDD LPP_unpleasant_ -MDD LPP_neutral_] *vs* [Healthy LPP_unpleasant_ -Healthy LPP_neutral_]. Numbered text to the right of the effect size bar graphs indicates *p*-values from independent samples *t*-tests comparing the LPP difference score (i.e. LPP_unpleasant_ - LPP_neutral_) between MDD and control participants. Results are formulated such that a negative Cohen’s *d* indicates support for *emotional context insensitivity*, and a positive Cohen’s *d* indicates support for *negative potentiation*. AR: Average reference; Localiser: collapsed localiser (Luck and Gaspelin, 2017); ROI: Region of interest (preregistered LPP channels Cz, CPz, Pz, CP1, and CP2);

Three additional exploratory non-parametric cluster-based permutation tests compared the difference scores (LPP_unpleasant_ - LPP_neutral_) between MDD participants and healthy controls using alternative pipelines (0.5Hz or 0.1Hz high-pass filters and referencing to linked 32 mastoids; and 0.1Hz high-pass filter using the common average reference). No significant 33 clusters were identified using these data-driven comparisons (all *p*s > 0.27)

## DISCUSSION

We performed a preregistered study to verify the presence of *emotional context insensitivity* in depression by comparing the LPP amplitude for unpleasant and neutral stimuli in MDD participants with age- and gender-matched healthy controls. Our preregistered data analysis plan found no difference in emotional reactivity between MDD participants and controls within the preregistered centroparietal region of interest and time interval. Further re-analysis of the data using 16 different EEG processing pipelines reported in the literature and five additional alternative pipelines also found no evidence of a difference in emotional reactivity between groups. Exploratory permutation analyses only revealed a significant difference between MDD participants and healthy individuals using the preregistered settings. This effect was in the hypothesised direction. The difference in amplitudes between unpleasant pictures relative to neutral pictures was lower for MDD participants compared to healthy controls, in agreement with the *emotional context insensitivity* model of depression. The exploratory results consisted of parietal EEG channels slightly posterior to the preregistered region of interest within a narrower post-stimulus time interval (662-755 ms). We found no evidence that emotion regulation strategy use differed between diagnostic groups, nor that strategy use or anxiety influenced the LPP in a passive view paradigm. Overall, our results provide no or weak evidence in support of *emotional context insensitivity* in our depressed sample and showed that quantification of the LPP in depression strongly depended on EEG processing pipeline parameters.

Evidence in support of emotional context insensitivity in depression arises from attenuated LPP responses to emotionally evocative stimuli (Hajcak and Foti, 2020), and from broader physiological research demonstrating reduced responses to both pleasant and unpleasant stimuli (Bylsma et al., 2008; Rottenberg et al., 2005; Rottenberg and Hindash, 2015). However, mixed effects have been reported in some EEG studies, including suggestions of reduced emotional reactivity to unpleasant stimuli in MDD relative to a healthy cohort (Foti et al., 2010; MacNamara et al., 2016), increased reactivity (Zhang et al., 2016), and no difference between unpleasant and neutral stimuli (Weinberg et al., 2016). The aim of the present study was therefore to obtain additional evidence for *emotional context insensitivity* in depression through a registered study (Nosek et al., 2018), replicating observations of reduced LPP to unpleasant stimuli in a large sample of adult MDD participants. We did not find evidence of a significant reduction in LPP amplitude for negatively-valenced stimuli using either our preregistered analysis plan or processing pipelines reported in literature. Similarly, no differences between groups were observed using a data-driven approach (i.e., cluster permutation testing), except for a statistically significant reduction in MDD LPP amplitudes using our preregistered EEG processing parameters (i.e., 0.5Hz high-pass filter and common average reference). This cluster significantly differentiated diagnostic groups within a narrower time window of the LPP (662-755 ms) and a slightly more posterior region of interest. Although these findings matched our *a priori* directional prediction (i.e., low LPP in the MDD group while viewing unpleasant images compared to healthy controls), they should be interpreted with caution. Only one exploratory analysis, out of a total of 26 variations reported in the present study, detected a significant LPP effect for the interaction contrast between groups for unpleasant and neutral images. While cluster-based permutation tests control the family-wise error rate (Maris and Oostenveld, 2007; Pernet et al., 2015), simulations conducted using similarly multidimensional neuroimaging data (obtained using fMRI) note that such procedures may have low sensitivity (i.e. true positive rate) (Noble et al., 2020). Furthermore, they should not be used to make inferences about the latency or location of effects (Sassenhagen and Draschkow, 2019). As such, these exploratory findings, which only marginally exceeded the significance threshold (*p* = 0.01), should not be used as conclusive evidence in favour of *emotional context insensitivity* in depression.

It is unclear why we did not observe more convincing evidence of attenuated LPP amplitudes in MDD sample. Potential reasons include the stimuli used to elicit the LPP, and the passive viewing study design. Stimuli selection differs markedly between studies, specifically with regards to the selection of aversive, unpleasant stimuli. These have included negatively-valenced scenes (MacNamara et al., 2016; Weinberg et al., 2016), affective and self-referential adjectives (Auerbach et al., 2015; Hilimire et al., 2015; Shestyuk and Deldin, 2010; Xie et al., 2018), and pictures of faces depicting sadness, anger, or fearful expressions (Foti et al., 2010; Zhang et al., 2016). The latter have been shown to produce a stronger amygdala response in comparison to matched fearful or threatening scenes (Hariri et al., 2002), which has important implications for the expected neurophysiological response measured using EEG (Mavratzakis et al., 2016). In addition to the type of stimulus, an important consideration is the context in which participants are instructed to interact with stimuli, either as part of a task, through active viewing of images with directed attention (e.g. using preselected emotional regulation strategies), or in a passive view paradigm. Hajcak et al. (2009) showed that instructing participants to attend to either neutral or arousing features in an image resulted in an attenuated LPP as compared to passive viewing. This agrees with research suggesting emotion regulation strategy use reduces LPP amplitudes (Dunning and Hajcak, 2009; Foti and Hajcak, 2008; Hajcak et al., 2007; Hajcak et al., 2006; Hajcak and Nieuwenhuis, 2006; Moser et al., 2006; Thiruchselvam et al., 2011; Uusberg et al., 2014; Zhang et al., 2014). Interestingly, in the present study we did not find supporting evidence that viewing strategies moderated outcomes in our dataset. An important caveat, however, is that we did not systematically instruct participants to adopt a particular strategy. Most participants in the current study reported no emotion regulation strategy use, thus limiting the ability to statistically infer the effects of strategy on LPP amplitudes.

A key outcome of the present study is that LPP findings may not necessarily generalise across stimulus processing steps and measurement contexts (Yarkoni, 2019). Sensitivity analyses performed by re-processing and operationalising the LPP using multiple parameter variations empirically demonstrate that relatively minor differences can impact effect estimates. Effect sizes were observed to change both in magnitude and direction depending on the EEG pipeline used to generate the LPP (Fig. 3). However, we note that there was considerable overlap in effect size intervals and that statistical results remained qualitatively unchanged regardless of processing methodology, i.e., none of the pipelines found a significant group by valence interaction effect. Based on these results it is plausible that mixed findings of emotional reactivity in the EEG studies described earlier may in part be caused by variations in pipeline processing steps and definitions of the LPP (see Table 3). Interpretation of findings within the context of the broader literature is therefore complicated by differences in the methodology, i.e., researcher degrees of freedom (Simmons et al., 2011), used to obtain the LPP. This provides a compelling argument for the development of a standardized EEG measure of emotional reactivity, encompassing the stimuli set, task protocol, EEG processing pipeline, and statistical analysis procedure used to assess the LPP. Though beyond the scope of the current manuscript, construction of such a tool could follow established methods to develop psychometric instruments of behaviour (Boateng et al., 2018).

### Limitations

A potential limitation to our study design is that depressed participants were allowed to be on stable ongoing antidepressant medications, which may have influenced LPP amplitudes (Kerestes et al., 2009). The impact of antidepressants on ERP components is unclear, having been found to both reduce (d’Ardhuy et al., 1999) or increase amplitudes (Ozgocmen et al., 2003; Sanz et al., 2001). We note that previous studies in depression have used similar recruitment strategies and have reported no differences in LPP component amplitudes between medicated and unmedicated participants (Auerbach et al., 2015; MacNamara et al., 2016; Webb et al., 2017).

Our stimuli set was restricted to negative and neutral pictures, omitting positive images. The reason for this was to reduce the total duration of the experiment session and minimise participant burden, as this study was part of a larger battery of cognitive tests. Given the weight of the evidence in the literature, we sought to replicate research in support of *emotional context insensitivity*, which uniquely predicts decreased reactivity in response to unpleasant stimuli.

## Conclusion

We investigated emotional reactivity in a large sample of MDD and age- and gender-matched controls to obtain additional evidence in support of *emotional context insensitivity* in depression. Contrary to our hypothesis, we did not find a statistical difference in emotional reactivity between groups using our preregistered analysis plan or the processing pipelines of several representative LPP studies in depression. Exploratory cluster analyses identified a single cluster, which was marginally more posterior and subtended a narrower time interval (660-755ms) than our preregistered LPP, that revealed a significant difference between MDD and control participants. However, as this cluster was the only significant finding of a total of 26 analysis variations, this only provides weak evidence in support of *emotional context insensitivity* in depression. Interestingly, EEG processing pipelines were found to affect both the magnitude and direction of the LPP comparison between MDD and control participants. These results suggest that methodological differences between studies (e.g., choice of affective stimuli, EEG processing pipeline, and analysis methods) may play an important role in observed heterogeneity of the cortical response to emotional stimuli in depression. This heterogeneity in EEG correlates of affective processing strongly indicates the need for a standardised EEG test and analysis pipeline for emotional reactivity.

## Data Availability

Participant consent was not obtained to make data publically available. However, SPSS syntax for mixed-effects repeated measures model statistical analyses, Inquisit 5 scripts to run the experimental protocol, and MATLAB scripts for EEG processing, cluster-based permutation tests, and plotting of figures, has been made available online at https://github.com/snikolin/MDDvsCTRL_IAPS.

## ACKNOWLEDGEMENTS

We would like to thank William Flannery, Liyi Tan and Yi Yin Tan for their contributions to EEG acquisitions.

