## Supplementary Materials for "Lack of Evidence for a Reduced Late Positive Potential in Major Depressive Disorder"

### EEG Correlates of Affective Processing in Major Depressive Disorder

#### SUPPLEMENTARY MATERIALS

##### IAPS Picture Set

A total of 56 pictures were selected from the International Affective Picture System (IAPS; Lang [1]). Of these, 28 were unpleasant pictures (e.g., threatening scenes, violence and weapons) and 28 were neutral pictures (e.g., neutral faces, household objects and landscapes). Pictures were additionally selected to be consistent with those used in previous EEG studies investigating LPP effects.

To eliminate potential distressing reactions associated with viewing unpleasant pictures, 10 research team members (some with prior history of depression) screened the pictures and by consensus removed images deemed likely to be too distressing. The final list of IAPS stimuli used in the experiment is presented in Table S1.

Normative arousal ratings were higher for unpleasant pictures (5.74 +- 0.84) compared to neutral pictures (2.98 +- 0.59) ($t(54) = 14.33$, $p < 0.001$), but lower in valence ratings (2.79 +- 0.59) compared to neutral pictures (5.02 +- 0.20) ($t(54) = 19.04$, $p < 0.001$).

**Supplementary Table S1. Normative ratings of the IAPS stimuli used in this study**

| | | Valence | | Arousal | | |
|---|---|---|---|---|---|---|
| IAPS List A | Description | Mean | SD | Mean | SD | Class |
| 6230 | AimedGun | 2.37 | 1.57 | 7.35 | 2.01 | Unpleasant |
| 2053 | Baby | 2.47 | 1.87 | 5.25 | 2.46 | Unpleasant |
| 9620 | Shipwreck | 2.70 | 1.64 | 6.11 | 2.10 | Unpleasant |
| 2745.1 | Shopping | 5.31 | 1.08 | 3.26 | 1.96 | Neutral |
| 7035 | Mug | 4.98 | 0.96 | 2.66 | 1.82 | Neutral |
| 2691 | Riot | 3.04 | 1.73 | 5.85 | 2.03 | Unpleasant |
| 7950 | Tissue | 4.94 | 1.21 | 2.28 | 1.81 | Neutral |
| 6821 | Gang | 2.38 | 1.72 | 6.29 | 2.02 | Unpleasant |
| 9520 | Kids | 2.46 | 1.61 | 5.41 | 2.27 | Unpleasant |
| 7150 | Umbrella | 4.72 | 1.00 | 2.61 | 1.76 | Neutral |
| 3300 | DisabledChild | 2.74 | 1.56 | 4.55 | 2.06 | Unpleasant |
| 2570 | Man | 4.78 | 1.24 | 2.76 | 1.92 | Neutral |
| 2688 | Hunters | 2.73 | 2.07 | 5.98 | 2.22 | Unpleasant |
| 2703 | SadChildren | 1.91 | 1.26 | 5.78 | 2.25 | Unpleasant |
| 2516 | ElderlyWoman | 4.90 | 1.43 | 3.50 | 1.88 | Neutral |
| 5532 | Mushrooms | 5.19 | 1.69 | 3.79 | 2.20 | Neutral |
| 1302 | Dog | 4.21 | 1.78 | 6.00 | 1.87 | Unpleasant |
| 2811 | Gun | 2.17 | 1.38 | 6.90 | 2.22 | Unpleasant |
| 2393 | FactoryWorker | 4.87 | 1.06 | 2.93 | 1.88 | Neutral |
| 2683 | War | 2.62 | 1.78 | 6.21 | 2.15 | Unpleasant |
| 7185 | AbstractArt | 4.97 | 0.87 | 2.64 | 2.04 | Neutral |
| 7034 | Hammer | 4.95 | 0.87 | 3.06 | 1.95 | Neutral |
| 2710 | DrugAddict | 2.52 | 1.69 | 5.46 | 2.29 | Unpleasant |
| 7100 | FireHydrant | 5.24 | 1.20 | 2.89 | 1.70 | Neutral |
| 5731 | Flowers | 5.39 | 1.58 | 2.74 | 1.95 | Neutral |
| 7037 | Trains | 4.81 | 1.12 | 3.71 | 2.08 | Neutral |
| 6260 | AimedGun | 2.44 | 1.54 | 6.93 | 1.93 | Unpleasant |
| 7050 | HairDryer | 4.93 | 0.81 | 2.75 | 1.80 | Neutral |
| 3220 | Hospital | 2.49 | 1.29 | 5.52 | 1.86 | Unpleasant |
| 7009 | Mug | 4.93 | 1.00 | 3.01 | 1.97 | Neutral |
| 2810 | Boy | 4.31 | 1.65 | 4.47 | 1.92 | Unpleasant |
| 2840 | Chess | 4.91 | 1.52 | 2.43 | 1.82 | Neutral |
| 7590 | Traffic | 4.75 | 1.55 | 3.80 | 2.13 | Neutral |
| 5535 | StillLife | 4.81 | 1.52 | 4.11 | 2.31 | Neutral |
| 3500 | Attack | 2.21 | 1.34 | 6.99 | 2.19 | Unpleasant |
| 2455 | SadGirls | 2.96 | 1.79 | 4.46 | 2.12 | Unpleasant |
| 7705 | Cabinet | 4.77 | 1.02 | 2.65 | 1.88 | Neutral |
| 7080 | Fork | 5.27 | 1.09 | 2.32 | 1.84 | Neutral |
| 2981 | DeerHead | 2.76 | 1.94 | 5.97 | 2.12 | Unpleasant |
| 2750 | Bum | 2.56 | 1.32 | 4.31 | 1.81 | Unpleasant |
| 6020 | ElectricChair | 3.41 | 1.98 | 5.58 | 2.01 | Unpleasant |
| 2200 | NeutFace | 4.79 | 1.38 | 3.18 | 2.17 | Neutral |
| 2141 | GrievingFem | 2.44 | 1.64 | 5.00 | 2.03 | Unpleasant |
| 1930 | Shark | 3.79 | 1.92 | 6.42 | 2.07 | Unpleasant |
| 6212 | Soldier | 2.19 | 1.49 | 6.01 | 2.44 | Unpleasant |
| 2191 | Farmer | 5.30 | 1.62 | 3.61 | 2.14 | Neutral |
| 2305 | Woman | 5.41 | 1.12 | 3.63 | 2.04 | Neutral |
| 7000 | RollingPin | 5.00 | 0.84 | 2.42 | 1.79 | Neutral |
| 6200 | AimedGun | 2.71 | 1.58 | 6.21 | 2.28 | Unpleasant |
| 7500 | Building | 5.33 | 1.44 | 3.26 | 2.18 | Neutral |
| 9920 | CarAccident | 2.50 | 1.52 | 5.76 | 1.96 | Unpleasant |
| 7004 | Spoon | 5.04 | 0.60 | 2.00 | 1.66 | Neutral |
| 7010 | Basket | 4.94 | 1.07 | 1.76 | 1.48 | Neutral |
| 7235 | Chair | 4.96 | 1.18 | 2.83 | 2.00 | Neutral |
| 2700 | Woman | 3.19 | 1.56 | 4.77 | 1.97 | Unpleasant |
| 6834 | Police | 2.91 | 1.73 | 6.28 | 1.90 | Unpleasant |

#### Emotion Regulation Questionnaire

An adapted emotion regulation questionnaire was used to identify which strategy, if any, participants used to moderate their emotional response while passively viewing pictures.

**Choose only one option:**

While viewing unpleasant pictures:

1. I did not use any strategy
2. I looked on the bright side
3. I thought about something else
4. I distanced myself from the picture
5. I did not do any of the above, I did…

#### Exploratory Cluster Channels

Channels from the significant cluster identified by the exploratory cluster-based permutation testing are displayed below. These are sorted as a proportion of the length of time they were identified relative to the total duration of the cluster time interval. Channels that were present for >75% of the cluster time interval were used in follow-up analyses.

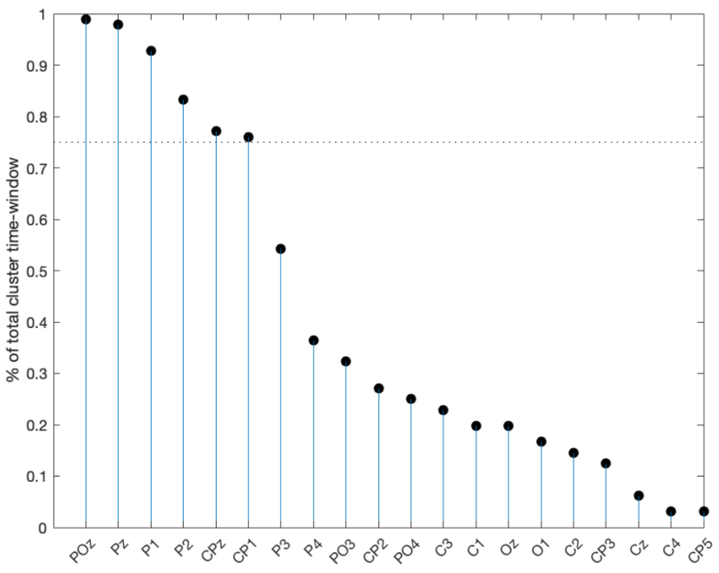

#### Effect of depressive symptoms on LPP amplitudes

Following the transdiagnostic, continuous approach espoused by the Research Domain Criteria Initiative (RDoC), we re-analysed LPP amplitude results using self-reported depressive symptoms. Mixed effects repeated measures models (MRMMs) included the repeated factor of Valence (unpleasant vs neutral) using an unstructured covariance matrix, in addition to the continuous variable of Depressive Symptoms (obtained from the DASS-21 depression subscore), and the Valence × Depressive Symptoms interaction. Subject was included as a random effect.

MRMM results showed a significant main effect of Valence (F = 8.8, $p < 0.001$). The main effect of Depressive Symptoms (F = 1.5, $p = 0.23$) and the Valence × Depressive Symptoms interaction (F = 0.1, $p = 0.79$) were not significant. These findings are highly similar to outcomes presented in the main body of the manuscript using categorical group assignment (i.e., MDD vs Control) instead of continuous depressive symptoms (i.e., DASS-21 depression subscore).

#### Secondary Analyses of Emotion Regulation Strategy and Anxiety

The LPP amplitude was compared using mixed effects repeated measures models (MRMMs) across Groups (MDD and controls) and Valence conditions (unpleasant and neutral), separately incorporating either Strategy (no strategy vs strategy) or Anxiety (from the DASS-21) as covariates. For preregistered LPP amplitudes the MRMM including Strategy as a covariate showed a significant main effect of Valence (F = 19.6, $p < 0.001$), but the main effects of Group (F = 0.5, $p$ = 0.50), the Group × Valence interaction (F = 0.3, $p$ = 0.61), and Strategy covariate (F = 0.2, $p$ = 0.67) were not significant (Fig. S1A). These results are similar to MRMM findings presented without Strategy as a covariate. Including Anxiety as a covariate likewise resulted in a significant main effect of Valence (F = 17.6, $p < 0.001$), but the main effects of Group (F = 0.1, $p$ = 0.77), the Group × Valence interaction (F = 0.4, $p$ = 0.51), and Anxiety covariate (F = 0.2, $p$ = 0.63) were not significant. There was no association between anxiety score and LPP amplitude for MDD participants while viewing unpleasant images (Fig. S1B).

Repeating the MRMMs above using the EEG channels and time interval identified in the exploratory cluster produced similar findings to results reported in the main body of the manuscript. The MRMM including Strategy as a covariate had a significant main effect of Valence (F = 9.6, $p < 0.001$), and Group × Valence interaction effect (F = 15.6, $p < 0.001$). Effects of Group (F = 0.6, $p$ = 0.43) and Strategy (F = 0.2, $p$ = 0.69) were not significant. Similarly, the MRMM including Anxiety as a covariate had a significant main effect of Valence (F = 8.5, $p$ = 0.01), and Group × Valence interaction effect (F = 17.5, $p < 0.001$). Effects of Group (F = 0.0, $p$ = 0.97) and Strategy (F = 2.6, $p$ = 0.11) were not significant.

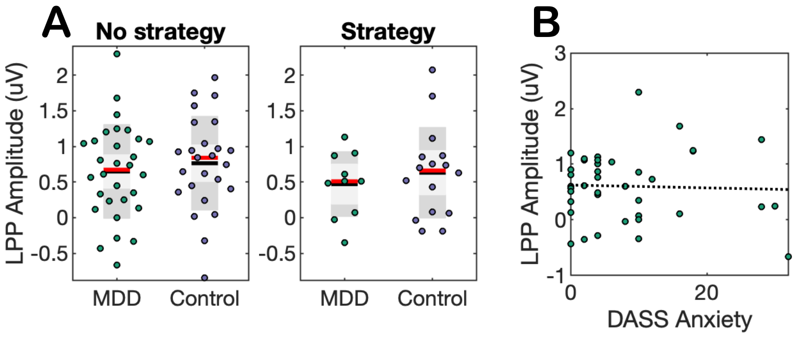


**Supplementary Figure S1. Effects of emotion regulation strategy and anxiety on LPP amplitude. A)** The LPP for the preregistered centroparietal cluster and time interval in both MDD participants and healthy controls who used an emotion regulation strategy or not. Black lines show the mean, red lines show the median, light grey shaded boxes indicate the standard deviation, and dark grey regions indicate the 95% confidence interval. **B)** Association between the anxiety subscore from the 21-item DASS and LPP amplitude for MDD participants while viewing unpleasant images. Dotted line shows the regression slope ($p > 0.05$; ns).

**Supplementary Table S2. Overview of statistical results for alternative analysis settings to calculate the LPP**

| | MDD LPP | | Control LPP | | MRMM Results | | |
|---|---|---|---|---|---|---|---|
| | Unpleasant | Neutral | Unpleasant | Neutral | Group | Valence | Group x Valence |
| *Originally submitted pipeline & analyses* | | | | | | | |
| Pre-registered pipeline | 0.61 ± 0.10 | 0.37 ± 0.10 | 0.71 ± 0.10 | 0.41 ± 0.10 | $F_{(1,79)} = 0.4$ $p = 0.54$ | $F_{(1,79)} = 19.6$ $p < 0.001$ | $F_{(1,79)} = 0.3$ $p = 0.61$ |
| Cluster-based permutation testing | 0.60 ± 0.13 | 0.67 ± 0.12 | 1.05 ± 0.13 | 0.46 ± 0.12 | $F_{(1,79)} = 0.6$ $p = 0.46$ | $F_{(1,79)} = 9.6$ $p = 0.003$ | $F_{(1,79)} = 15.6$ $p < 0.001$ |
| *Reviewer-suggested pipeline & analyses* | | | | | | | |
| Alternative #1 | 0.49 ± 0.26 | -0.13 ± 0.21 | 0.51 ± 0.25 | -0.02 ± 0.21 | $F_{(1,79)} = 0.1$ $p = 0.82$ | $F_{(1,79)} = 17.5$ $p < 0.001$ | $F_{(1,79)} = 0.0$ $p = 0.75$ |
| Alternative #2 | 1.09 ± 0.59 | -0.64 ± 0.57 | 1.33 ± 0.57 | 0.04 ± 0.55 | $F_{(1,78)} = 0.4$ $p = 0.53$ | $F_{(1,78)} = 21.3$ $p < 0.001$ | $F_{(1,78)} = 0.5$ $p = 0.50$ |
| Alternative #3 | 2.40 ± 0.53 | 0.48 ± 0.51 | 3.47 ± 0.52 | 1.73 ± 0.49 | $F_{(1,78)} = 3.1$ $p = 0.09$ | $F_{(1,78)} = 40.2$ $p < 0.001$ | $F_{(1,78)} = 0.1$ $p = 0.75$ |
| Alternative #4 | 1.42 ± 0.24 | 0.86 ± 0.22 | 1.76 ± 0.24 | 1.17 ± 0.21 | $F_{(1,78)} = 1.2$ $p = 0.28$ | $F_{(1,78)} = 21.7$ $p < 0.001$ | $F_{(1,78)} = 0.0$ $p = 0.89$ |
| Alternative #5 | 2.73 ± 0.40 | 1.98 ± 0.37 | 3.90 ± 0.39 | 2.86 ± 0.36 | $F_{(1,78)} = 4.0$ $p = 0.05$ | $F_{(1,78)} = 36.2$ $p < 0.001$ | $F_{(1,78)} = 1.0$ $p = 0.33$ |
| Representative LPP studies in depression | | | | | | | |
| Foti 2010; Macnamara 2019 | 0.57 ± 0.52 | -1.09 ± 0.46 | 0.02 ± 0.51 | -1.13 ± 0.45 | $F_{(1,78)} = 0.2$ $p = 0.64$ | $F_{(1,78)} = 27.2$ $p < 0.001$ | $F_{(1,78)} = 0.9$ $p = 0.35$ |
| Shestyuk 2010 | -1.89 ± 0.72 | -3.03 ± 0.60 | -2.77 ± 0.72 | -3.62 ± 0.60 | $F_{(1,78)} = 0.8$ $p = 0.38$ | $F_{(1,78)} = 5.5$ $p = 0.02$ | $F_{(1,78)} = 0.1$ $p = 0.74$ |
| Hilimire 2015 | 2.30 ± 0.34 | 1.69 ± 0.33 | 3.51 ± 0.34 | 2.32 ± 0.33 | $F_{(1,76)} = 4.4$ $p = 0.04$ | $F_{(1,76)} = 29.1$ $p < 0.001$ | $F_{(1,76)} = 3.0$ $p = 0.09$ |
| Auerbach 2015 | -0.66 ± 0.30 | -1.15 ± 0.28 | -1.08 ± 0.30 | -1.34 ± 0.28 | $F_{(1,78)} = 0.7$ $p = 0.42$ | $F_{(1,78)} = 6.0$ $p = 0.02$ | $F_{(1,78)} = 0.5$ $p = 0.48$ |
| Weinberg 2016 | -1.21 ± 0.53 | -0.87 ± 0.48 | -0.90 ± 0.52 | -0.73 ± 0.47 | $F_{(1,78)} = 0.0$ $p = 0.90$ | $F_{(1,78)} = 35.0$ $p < 0.001$ | $F_{(1,78)} = 0.5$ $p = 0.48$ |
| Zhang 2016 | 0.54 ± 0.56 | -0.94 ± 0.52 | 0.30 ± 0.54 | -0.73 ± 0.50 | $F_{(1,75)} = 0.0$ $p = 0.99$ | $F_{(1,75)} = 19.0$ $p < 0.001$ | $F_{(1,75)} = 0.6$ $p = 0.44$ |
| Stange 2017a; Stange 2017b | 0.71 ± 0.56 | -1.38 ± 0.50 | 0.06 ± 0.54 | -1.37 ± 0.49 | $F_{(1,78)} = 0.2$ $p = 0.63$ | $F_{(1,78)} = 29.6$ $p < 0.001$ | $F_{(1,78)} = 1.1$ $p = 0.31$ |
| Burkhouse 2017 | 1.06 ± 0.39 | -0.02 ± 0.37 | 1.58 ± 0.38 | 0.53 ± 0.36 | $F_{(1,78)} = 1.2$ $p = 0.28$ | $F_{(1,78)} = 33.2$ $p < 0.001$ | $F_{(1,78)} = 9.9$ $p = 0.94$ |

| | | | | | | | |
|---|---|---|---|---|---|---|---|
| Webb 2017 | -0.76 ± 0.29 | -1.35 ± 0.28 | -1.21 ± 0.29 | -1.49 ± 0.28 | $F_{(1,78)} = 0.6$ $p = 0.43$ | $F_{(1,78)} = 7.2$ $p = 0.01$ | $F_{(1,78)} = 1.0$ $p = 0.33$ |
| Xie 2018 | 1.54 ± 0.57 | -0.22 ± 0.46 | 2.01 ± 0.56 | 0.29 ± 0.45 | $F_{(1,79)} = 0.58$ $p = 0.45$ | $F_{(1,79)} = 26.0$ $p < 0.001$ | $F_{(1,79)} = 0.0$ $p = 0.95$ |
| Buchheim 2018 | -0.42 ± 0.20 | -0.88 ± 0.20 | -1.27 ± 0.20 | -1.42 ± 0.21 | $F_{(1,79)} = 7.1$ $p = 0.01$ | $F_{(1,79)} = 6.3$ $p = 0.01$ | $F_{(1,79)} = 1.7$ $p = 0.20$ |
| Grunewald 2019 | 1.12 ± 0.17 | 0.79 ± 0.15 | 1.52 ± 0.17 | -1.05 ± 0.15 | $F_{(1,79)} = 2.8$ $p = 0.10$ | $F_{(1,79)} = 13.9$ $p < 0.001$ | $F_{(1,79)} = 0.4$ $p = 0.52$ |
| Benau 2019 | 1.21 ± 0.58 | -1.01 ± 0.49 | 1.09 ± 0.56 | -0.66 ± 0.48 | $F_{(1,78)} = 0.0$ $p = 0.86$ | $F_{(1,78)} = 36.0$ $p < 0.001$ | $F_{(1,78)} = 0.5$ $p = 0.48$ |
| Macnamara 2019 | 1.49 ± 0.55 | -0.79 ± 0.46 | 0.96 ± 0.55 | -0.87 ± 0.45 | $F_{(1,79)} = 0.3$ $p = 0.58$ | $F_{(1,79)} = 20.8$ $p < 0.001$ | $F_{(1,79)} = 0.3$ $p = 0.61$ |
| Trotti 2020 | -0.18 ± 0.25 | -0.67 ± 0.22 | -0.26 ± 0.25 | -0.52 ± 0.22 | $F_{(1,78)} = 0.0$ $p = 0.91$ | $F_{(1,78)} = 9.9$ $p < 0.001$ | $F_{(1,78)} = 0.9$ $p = 0.34$ |
| Whalen 2020 | -0.46 ± 0.54 | -0.67 ± 0.23 | -0.67 ± 0.54 | -0.59 ± 0.23 | $F_{(1,70.4)} = 3.2$ $p = 0.08$ | $F_{(1,76.5)} = 2.4$ $p = 0.13$ | $F_{(1, 76.5)} = 1.2$ $p = 0.28$ |
